# Assessing Folic Acid Supplementation Adherence Among Pregnant Women in LMICs and its Impact on Neural Tube Defect Incidence: A Systematic Review Protocol

**DOI:** 10.1101/2023.09.26.23296200

**Authors:** Jeremiah Oluwatomi Itodo Daniel, Abdulmuizz Oluwatomi Sulaiman, Chiwendu Isiakpona, Joanne Igoli, Chukwunonso Emmanuel Chukwumaeze, Eghosa Morgan, Olatomiwa Olukoya

## Abstract

**Introduction:** Neural tube defects (NTDs) are treatable but incurable birth defects, causing permanent disability and potential mortality if neglected. Despite its global significance, there has not been any systematic review exploring the link between folic acid adherence and NTD incidence in low-and-middle-income countries (LMICs), where its impact is disproportionately felt. This systematic review primarily aims to analyze the impact of folic acid supplementation adherence among expectant mothers on the incidence of NTDs in LMICs.

**Methods and Analysis:** Following the PRISMA guidelines, we will conduct a comprehensive search of original research articles and observational studies using the electronic databases of PubMed, Scopus, Medline, and Embase from inception till date. Our search comprises keywords related to our PICO framework and objectives. We will analyze data using tables and diagrams for qualitative presentation and conduct a random-effect Bayesian meta-analysis when applicable.

Two reviewers will screen titles and abstracts independently, conduct full-text reviews, and extract study data. In case of disagreements, a third reviewer will resolve them. We will present study details and assess bias risk. We will also assess the overall certainty of evidence using the Grading of Recommendations Assessment, Development, and Evaluation framework.

**Ethics and Dissemination:** Ethical approval is unnecessary as this study involves a systematic review of published studies. We aim to publish our findings in a peer-reviewed journal.

## Background and Rationale

Neural tube defects (NTDs) remain a dreadful birth defect, with spina bifida and anencephaly among the most common types. While anencephaly is lethal and incompatible with life, open spina bifida, such as myelomeningocele, can be treated but not cured. This can lead to severe lifelong disability and, if untreated, death. (1)

Physical and psychological issues arise with the permanent disability from Spina bifida, necessitating continuous surgical and medical care. Globally, an estimated 300,000 babies are born with NTDs yearly. In low and middle-income countries (LMICs), about 60,000 pregnancies affected by NTDs are euthanized following prenatal diagnosis, while another 60,000 result in stillbirths. (1) Salari et al., 2021 estimate the average prevalence rate of spina bifida and anencephaly as 20 cases per 100,000 live births worldwide. (2) There exists a paucity of data on NTDs in the LMICs; however, Yacob et al. (2021) showed that countries with the highest years lived in disability and death rates due to NTDs were the lowest gross domestic product (GDP) quartile. (3)

Surgical treatment for neural tube defects in LMICs is riddled with complexity, high costs, and the limits of a low neurosurgical and multidisciplinary workforce. (4–6) Public health programs educating women of reproductive age of folic acid importance are almost absent, hence the rising trend of NTDs.

A significant proportion of pregnant women globally do not attain the recommended daily intake of 0.4mg of Folic acid for six months (7). Folate deficiency places these pregnant women at a higher risk of having newborns with NTDs.

However, among other factors, such as forgetting to take folate, that may hinder the broader adoption of folic acid supplementation, pregnant women in LMICs often face numerous health challenges. These include further nutrient deficiencies, inadequate healthcare access, and poor or non-existent prenatal care, which prevent the early diagnosis or prevention of NTDs in their newborns(8).

This systematic review aims to critically evaluate the evidence of folic acid supplementation adherence among pregnant women in LMICs and its effect on NTD incidence. We define adherence as the degree to which a patient complies with treatment recommendations made by a healthcare professional.

By conducting this review, we seek to address significant gaps in the literature and inform public health policies and interventions targeted at improving maternal and child health outcomes. The significance of this systematic review lies in its potential to provide valuable insights into the overall effectiveness of this intervention and its potential to mitigate adverse pregnancy outcomes.

## Research Questions

1. What is the current folic acid supplementation adherence level among pregnant women in LMICs?
2. How does compliance with folic acid supplementation among pregnant women affect the incidence of neural tube defects in LMICs?
3. What predictors influence adherence to folic acid supplementation among pregnant women in LMICs?

## Objectives and Outcome Measures

### Specific Objectives

- To systematically review and analyze studies assessing folic acid supplementation adherence in pregnant women across LMICs.
- To examine the impact of folic acid supplementation adherence on the incidence of neural tube defects in newborns within LMICs.
- To identify factors influencing adherence to folic acid supplementation among pregnant women in LMICs.

## Population

Pregnant women living in LMICs.

## Intervention/Exposure

Folic acid supplementation during pregnancy.

## Comparator

Pregnant women who take folic acid supplementation compared to those who do not.

## Outcome

### Primary Outcomes

- Adherence to Folic acid supplementation during pregnancy
- Incidence of neural tube defects

## Methods

### Literature Search - Search Strategy

This systematic review will follow the Preferred Reporting Items for Systematic Review and Meta-Analysis Protocols (PRISMA-P) guidelines. We will perform a comprehensive literature search primarily on PubMed, SCOPUS, Medline, and Embase databases from inception till date.

We will search the titles/abstracts/keywords on these databases as follows:

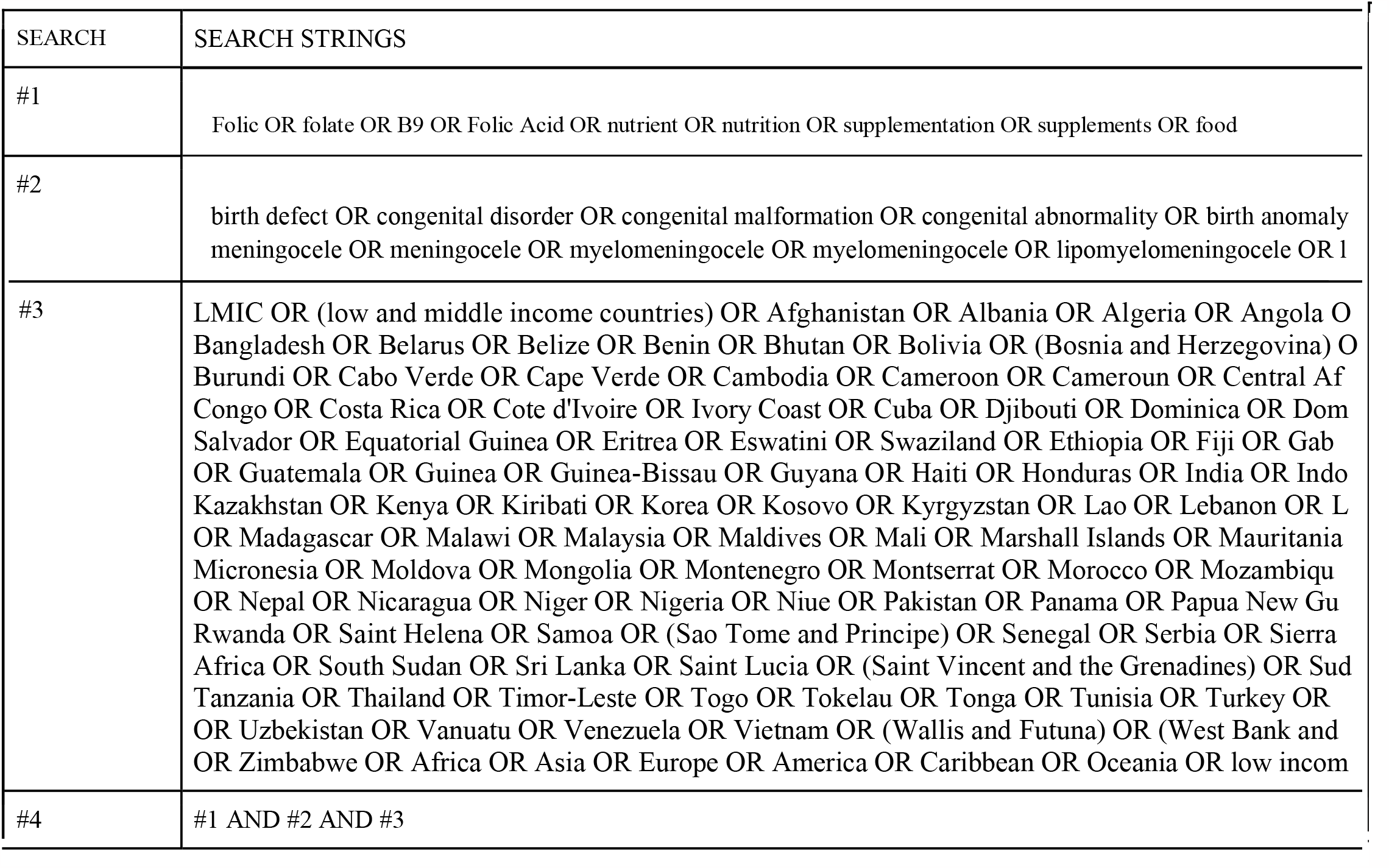

Following title and abstract screening, the reference list of full-text papers for inclusion will be searched for additional studies to include.

### Inclusion Criteria

A study will be included if it meets the following criteria:

1. Observational studies on the compliance of folic acid supplementation on pregnant women in LMICs.
2. Original research articles on folic acid supplementation adherence in pregnant women and its association with NTDs within the LMICs..
3. Studies published in English or with available English translations.

### Exclusion Criteria

A study will be excluded if it meets these criteria:

1. Review articles, expert opinions, case reports, abstract-only articles, conference proceedings, meta-analyses, and letters to the editor.
2. Studies that do not specifically assess folic acid supplementation adherence without information on the incidence of NTDs.
3. Non-human subjects.
4. Reports on non-dietary sources of folic acid.
5. Subjects with a genetic predisposition to develop NTDs or syndromic manifestations of NTDs (i.e., TORCHES syndrome infection).

## Data Management, Selection of Sources of Evidence, and Data Charting

We will devise a comprehensive search strategy using keywords and MeSH terms related to the topic from electronic databases such as PubMed, Ovid MEDLINE, Scopus, and Embase. We will exclude literature search duplicates. Using Rayyan software, initial screening will involve the assessment of titles and abstracts to exclude irrelevant or unrelated studies. Subsequently, full-text articles of potentially eligible studies will be retrieved for detailed evaluation.

Two reviewers will independently screen a set of full-text articles and perform data extraction using a predesigned proforma. Any reviewer disagreements will be resolved through discussion or consultation with a third reviewer if necessary.

The process will be documented in a transparent manner using a Preferred Reporting Items for Systematic Reviews and Meta-Analyses (PRISMA) flow diagram. The final selection of studies will form the basis for the systematic review’s subsequent data extraction and analysis phases.

### Data Charting

This will be done on Google Sheets (Google, Mountain View, CA, USA) to ensure all reviewers’ data is homogeneously extracted. The following data will be extracted:

- Title
- Author
- Year of publication
- Study design: prospective versus retrospective; cohort study versus case-controlled study versus others
- Country of study
  - Study population
- Age (mean + SD/median (IQR))
- Sex (number of males/number of females)
- Event ratios

### Data Extraction

Data will be extracted using a standardized form, including study characteristics, participant details, adherence measurement methods, outcomes, and factors influencing adherence. The quality of included studies will be assessed using appropriate tools, such as the Newcastle-Ottawa Scale for observational studies and the Cochrane risk of bias tool for randomized controlled trials.

### Data Analysis

The analysis will be conducted using the Revman software v.5.4. The degree of heterogeneity among the included studies will be assessed using the I^2^ statistic. A value exceeding 50% for the I^2^ statistic will indicate significant heterogeneity. Statistical significance will be determined by P<0.05.

### Data Synthesis

Data will be synthesized qualitatively through narrative synthesis. If sufficient data and homogeneity exist, a meta-analysis will be conducted using a random-effects model. Subgroup and sensitivity analyses will be performed to explore heterogeneity and assess the robustness of the results.

## Subgroup Analysis

If we find any significant heterogeneity, we will conduct a subgroup analysis based on geographic region, study design, and adherence measurement methods. Sensitivity analyses will be performed to test the robustness of results by excluding studies with a high risk of bias or low methodological quality. Subgroup analyses will be performed based on study design, geographic location, folic acid supplementation initiation timing, and the presence or absence of TORCHES syndrome infection in the study population.

## Evaluation of Heterogeneity

We anticipate that studies with different observational designs may exhibit varying degrees of heterogeneity. We will utilize statistical tests such as Cochran’s Q and I^2^ statistics to quantify heterogeneity across studies. A significant Q test or a high I^2^ value suggests substantial heterogeneity.

## Strength of Body Evidence

The quality of evidence will be assessed using the Grading of Recommendations Assessment, Development, and Evaluation (GRADE) approach, considering factors such as study design, risk of bias, consistency, precision, and publication bias. Two reviewers will assess the certainty of evidence for each outcome independently. A third reviewer will be available to resolve conflicts in the event of conflicts. We will document each comparison by outcome, including explanations for all decisions on the GRADE Summary of Findings tables. We will exclude studies with a high risk of bias or those with methodological limitations.

## Risk of Bias (Quality) Assessment

Two independent reviewers will meticulously assess within-study and cross-study biases for each outcome measure. The quality of observational studies will be appraised using the Newcastle-Ottawa Scale (NOS). At the same time, the risk of bias in randomized controlled trials (RCTs) will be evaluated through The Cochrane Collaboration risk of bias tool. Only studies that achieve an NOS score of ≥4 will be deemed to possess a low risk of bias and, therefore, considered eligible for inclusion in the systematic review and subsequent meta-analysis.

To evaluate the impact of potential publication bias on the outcomes, funnel plots and statistical tests (e.g., Egger’s test) will be employed. Studies with small sample sizes or those that may not have been published due to non-significant results will be considered.

## Measurement of Effect

We will analyze data from each group: pregnant women who adhered to Folic acid supplementation versus those who did not. The odds ratios for each group will be calculated to determine the effect on the incidence of NTDs. We will compute the mean or standard mean difference for continuous, numerical, and categorical variables. Additionally, multivariate analyses will be conducted to control for potential confounding factors, such as risk of NTD recurrence, dietary intake of folate-rich foods, and patient characteristics.

## Study Limitations

Despite our comprehensive research methodology, notable limitations warrant consideration. First, publication bias is inevitable, as studies with statistically significant findings are more likely to be published than studies reporting less significant outcomes.

Moreover, the possibility of language bias exists, as our review exclusively includes English-language publications, potentially omitting relevant research published in other languages. Additionally, it is pertinent to acknowledge the potential for bias within the studies included in this systematic review, which could potentially influence our study’s overall outcomes and findings.

## Supporting information

Supplemental Tables - search results

## Data Availability

All data found in this research protocol are available at PubMed:https://pubmed.ncbi.nlm.nih.gov/
Scopus:https://www.scopus.com/
Ovid Medline:https://ovidsp.ovid.com/
Embase: https://www.embase.com/landing?status=grey

## Acknowledgments

JOID- Conceptualization, Methodology, Visualization, Writing, and Proofreading

All authors provided expert opinions, proofread, and edited the manuscript for final publication.

## Ethics and Dissemination

Since the review involves the analysis of existing studies, it does not involve direct contact with human participants. We plan to submit this systematic review for publication in a reputable peer-reviewed public health or maternal and child health journal. Also, we will present the findings at relevant academic conferences, both nationally and internationally, to reach a diverse audience of researchers and practitioners in the maternal and child health field.

## References

1. https://www.cdc.gov/mmwr/preview/mmwrhtml/mm6401a2.htm?s_cid=mm6401a2_w

1. Kancherla V, Botto LD, Rowe LA, Shlobin NA, Caceres A, Arynchyna-Smith A, et al. Preventing birth defects, saving lives, and promoting health equity: an urgent call to action for universal mandatory food fortification with folic acid. Lancet Glob Health. 2022 Jul 1;10(7):e1053–7.

2. Salari N, Fatahi B, Fatahian R, Mohammadi P, Rahmani A, Darvishi N, et al. Global prevalence of congenital anencephaly: a comprehensive systematic review and meta-analysis. Reprod Health. 2022 Oct 17;19(1):201.

3. Yacob A, Carr CJ, Foote J, Scullen T, Werner C, Mathkour M, et al. The global burden of neural tube defects and disparities in neurosurgical care. World Neurosurg. 2021;149:e803–20.

4. Park KB, Johnson WD, Dempsey RJ. Global Neurosurgery: The Unmet Need. World Neurosurg. 2016 Apr 1;88:32–5.

5. Punchak M, Mukhopadhyay S, Sachdev S, Hung YC, Peeters S, Rattani A, et al. Neurosurgical Care: Availability and Access in Low-Income and Middle-Income Countries. World Neurosurg. 2018 Apr 1;112:e240–54.

6. Updated Estimates of Neural Tube Defects Prevented by Mandatory Folic Acid Fortification — United States, 1995–2011 [Internet]. [cited 2023 Mar 4]. Available from: https://www.cdc.gov/mmwr/preview/mmwrhtml/mm6401a2.htm?s_cid=mm6401a2_w

7. Martinez H, Benavides-Lara A, Arynchyna-Smith A, Ghotme KA, Arabi M, Arynchyn A. Global strategies for the prevention of neural tube defects through the improvement of folate status in women of reproductive age. Childs Nerv Syst. 2023 Jul;39(7):1719–36.

8. Shlobin NA, Roach JT, Kancherla V, Caceres A, Ocal E, Ghotme KA, et al. The role of neurosurgeons in global public health: the case of folic acid fortification of staple foods to prevent spina bifida. J Neurosurg Pediatr. 2023 Jan 1;31(1):8–15.

