## Supplemental Tables - search results for "Assessing Folic Acid Supplementation Adherence Among Pregnant Women in LMICs and its Impact on Neural Tube Defect Incidence: A Systematic Review Protocol"

### **Search Strategy Tables**

DATABASE(S) SEARCHED: PubMed, Scopus, Ovid Medline, and Embase

**PubMed**

| SEARCH | SEARCH STRATEGY | RESULTS | DATE OF SEARCH | DATE RANGE |
| --- | --- | --- | --- | --- |
| #1 | Folic OR folate OR B9 OR Folic Acid OR nutrient OR nutrition OR supplementation OR supplements OR food | 2,593,002 | 24th September 2023 | From Inception till September 24, 2023 |
| #2 | birth defect OR congenital disorder OR congenital malformation OR congenital abnormality OR birth anomaly OR neural tube OR NTD OR spina bifida OR meningocele OR meningocele OR myelomeningocele OR myelomeningocele OR lipomyelomeningocele OR lipomyelomeningocele OR anencephaly | 1,445,832 | 24th September 2023 | From Inception till September 24, 2023 |
| #3 | LMIC OR (low and middle income countries) OR Afghanistan OR Albania OR Algeria OR Angola OR Argentina OR Armenia OR Azerbaijan OR Bangladesh OR Belarus OR Belize OR Benin OR Bhutan OR Bolivia OR (Bosnia and Herzegovina) OR Botswana OR Brazil OR Burkina Faso OR Burundi OR Cabo Verde OR Cape Verde OR Cambodia OR Cameroon OR Cameroun OR Central African Republic OR Chad OR China OR Colombia OR Congo OR Costa Rica OR Cote d'Ivoire OR Ivory Coast OR Cuba OR Djibouti OR Dominica OR Dominican Republic OR Ecuador OR Egypt OR El Salvador OR Equatorial Guinea OR Eritrea OR Eswatini OR Swaziland OR Ethiopia OR Fiji OR Gabon OR Gambia OR Georgia OR Ghana OR Grenada OR Guatemala OR Guinea OR Guinea-Bissau OR Guyana OR Haiti OR Honduras OR India OR Indonesia OR Iran OR Iraq OR Jamaica OR Jordan OR Kazakhstan OR Kenya OR Kiribati OR Korea OR Kosovo OR Kyrgyzstan OR Lao OR Lebanon OR Lesotho OR Liberia OR Libya OR North Macedonia OR Madagascar OR Malawi OR Malaysia OR Maldives OR Mali OR Marshall Islands OR Mauritania OR Mauritius OR Mexico OR Micronesia OR Micronesia OR Moldova OR Mongolia OR Montenegro OR Montserrat OR Morocco OR Mozambique OR Myanmar OR Burma OR Namibia OR Nauru OR Nepal OR Nicaragua OR Niger OR Nigeria OR Niue OR Pakistan OR Panama OR Papua New Guinea OR Paraguay OR Peru OR Philippines OR Rwanda OR Saint Helena OR Samoa OR (Sao Tome and Principe) OR Senegal OR Serbia OR Sierra Leone OR Solomon Islands OR Somalia OR South Africa OR South Sudan OR Sri Lanka OR Saint Lucia OR (Saint Vincent and the Grenadines) OR Sudan OR Suriname OR Syria OR Tajikistan OR Tanzania OR Thailand OR Timor-Leste OR Togo OR Tokelau OR Tonga OR Tunisia OR Turkey OR Turkmenistan OR Tuvalu OR Uganda OR Ukraine OR Uzbekistan OR Vanuatu OR Venezuela OR Vietnam OR (Wallis and Futuna) OR (West Bank and Gaza Strip) OR Palestine OR Yemen OR Zambia OR Zimbabwe OR Africa OR Asia OR Europe OR America OR Caribbean OR Oceania OR low income OR middle income | 10,866,010 | 24th September 2023 | From Inception till September 24, 2023 |
| #4 | #1 AND #2 AND #3 | 22,927 | 24th September 2023 | From Inception till September 24, 2023 |

**Scopus**

| SEARCH | SEARCH STRATEGY | RESULTS | DATE OF SEARCH | DATE RANGE |
| --- | --- | --- | --- | --- |
| #1 | Folic OR folate OR B9 OR Folic Acid OR nutrient OR nutrition OR supplementation OR supplements OR food | 93,056 | 24th September 2023 | From Inception till September 24, 2023 |
| #2 | birth defect OR congenital disorder OR congenital malformation OR congenital abnormality OR birth anomaly OR neural tube OR NTD OR spina bifida OR meningocele OR meningocele OR myelomeningocele OR myelomeningocele OR lipomyelomeningocele OR lipomyelomeningocele OR anencephaly | 928 | 24th September 2023 | From Inception till September 24, 2023 |
| #3 | LMIC OR (low and middle income countries) OR Africa OR low income OR middle income | 202,419 | 24th September 2023 | From Inception till September 24, 2023 |
| #4 | #1 AND #2 AND #3 | 314 | 24th September 2023 | From Inception till September 24, 2023 |

**MEDLINE (via Ovid)**

| SEARCH | SEARCH STRATEGY | RESULTS | DATE OF SEARCH | DATE RANGE |
| --- | --- | --- | --- | --- |
| #1 | Folic.ab,kw,ti. OR folate.ab,kw,ti. OR B9.ab,kw,ti. OR nutrient.ab,kw,ti. OR nutrition.ab,kw,ti. OR fortification.ab,kw,ti. OR policy.ab,kw,ti. OR policies.ab,kw,ti. OR supplement*.ab,kw,ti. OR food.ab,kw,ti. | 1550697 | 21st September 2023 | From Inception till September 21, 2023 |
| #2 | "birth defect*".ab,kw,ti. OR "congenital disorder*".ab,kw,ti. OR "congenital malformation*".ab,kw,ti. OR "congenital abnormalit*".ab,kw,ti. OR "birth anomal*".ab,kw,ti. OR neural tube.ab,kw,ti. OR NTD.ab,kw,ti. OR spina bifida.ab,kw,ti. OR meningocele.ab,kw,ti. OR meningocoele.ab,kw,ti. OR myelomeningocele.ab,kw,ti. OR myelomeningocoele.ab,kw,ti. OR lipomyelomeningocele.ab,kw,ti. OR lipomyelomeningocoele.ab,kw,ti. OR anencephaly.ab,kw,ti. | 70286 | 21st September 2023 | From Inception till September 21, 2023 |
| #3 | LMIC.ab,kw,ti OR (low and middle income countr*).ab,kw,ti OR Afghanistan.ab,kw,ti OR Albania.ab,kw,ti OR Algeria.ab,kw,ti OR Angola.ab,kw,ti OR Argentina.ab,kw,ti OR Armenia.ab,kw,ti OR Azerbaijan.ab,kw,ti OR Bangladesh.ab,kw,ti OR Belarus.ab,kw,ti OR Belize.ab,kw,ti OR Benin.ab,kw,ti OR Bhutan.ab,kw,ti OR Bolivia.ab,kw,ti OR (Bosnia and Herzegovina).ab,kw,ti OR Botswana.ab,kw,ti OR Brazil.ab,kw,ti OR Burkina Faso.ab,kw,ti OR Burundi.ab,kw,ti OR Cabo Verde.ab,kw,ti OR Cape Verde.ab,kw,ti OR Cambodia.ab,kw,ti OR Cameroon.ab,kw,ti OR Cameroun.ab,kw,ti OR Central African Republic.ab,kw,ti OR Chad.ab,kw,ti OR China.ab,kw,ti OR Colombia.ab,kw,ti OR Congo.ab,kw,ti OR Costa Rica.ab,kw,ti OR Cote d'Ivoire.ab,kw,ti OR Ivory Coast.ab,kw,ti OR Cuba.ab,kw,ti OR Djibouti.ab,kw,ti OR Dominica.ab,kw,ti OR Dominican Republic.ab,kw,ti OR Ecuador.ab,kw,ti OR Egypt.ab,kw,ti OR El Salvador.ab,kw,ti OR Equatorial Guinea.ab,kw,ti OR Eritrea.ab,kw,ti OR Eswatini.ab,kw,ti OR Swaziland.ab,kw,ti OR Ethiopia.ab,kw,ti OR Fiji.ab,kw,ti OR Gabon.ab,kw,ti OR Gambia.ab,kw,ti OR Georgia.ab,kw,ti OR Ghana.ab,kw,ti OR Grenada.ab,kw,ti OR Guatemala.ab,kw,ti OR Guinea.ab,kw,ti OR Guinea-Bissau.ab,kw,ti OR Guyana.ab,kw,ti OR Haiti.ab,kw,ti OR Honduras.ab,kw,ti OR India.ab,kw,ti OR Indonesia.ab,kw,ti OR Iran.ab,kw,ti OR Iraq.ab,kw,ti OR Jamaica.ab,kw,ti OR Jordan.ab,kw,ti OR Kazakhstan.ab,kw,ti OR Kenya.ab,kw,ti OR Kiribati.ab,kw,ti OR Korea.ab,kw,ti OR Kosovo.ab,kw,ti OR Kyrgyzstan.ab,kw,ti OR "Lao*".ab,kw,ti OR Lebanon.ab,kw,ti OR Lesotho.ab,kw,ti OR Liberia.ab,kw,ti OR Libya.ab,kw,ti OR North Macedonia.ab,kw,ti OR Madagascar.ab,kw,ti OR Malawi.ab,kw,ti OR Malaysia.ab,kw,ti OR Maldives.ab,kw,ti OR Mali.ab,kw,ti OR Marshall Islands.ab,kw,ti OR Mauritania.ab,kw,ti OR Mauritius.ab,kw,ti OR Mexico.ab,kw,ti OR Micronesia.ab,kw,ti OR Micronesia.ab,kw,ti OR Moldova.ab,kw,ti OR Mongolia.ab,kw,ti OR Montenegro.ab,kw,ti OR Montserrat.ab,kw,ti OR Morocco.ab,kw,ti OR Mozambique.ab,kw,ti OR Myanmar.ab,kw,ti OR Burma.ab,kw,ti OR Namibia.ab,kw,ti OR Nauru.ab,kw,ti OR Nepal.ab,kw,ti OR Nicaragua.ab,kw,ti OR Niger.ab,kw,ti OR Nigeria.ab,kw,ti OR Niue.ab,kw,ti OR Pakistan.ab,kw,ti OR Panama.ab,kw,ti OR Papua New Guinea.ab,kw,ti OR Paraguay.ab,kw,ti OR Peru.ab,kw,ti OR Philippines.ab,kw,ti OR Rwanda.ab,kw,ti OR Saint Helena.ab,kw,ti OR Samoa.ab,kw,ti OR (Sao Tome and Principe).ab,kw,ti OR Senegal.ab,kw,ti OR Serbia.ab,kw,ti OR Sierra Leone.ab,kw,ti OR Solomon Islands.ab,kw,ti OR Somalia.ab,kw,ti OR South Africa.ab,kw,ti OR South Sudan.ab,kw,ti OR Sri Lanka.ab,kw,ti OR Saint Lucia.ab,kw,ti OR (Saint Vincent and the Grenadines).ab,kw,ti OR Sudan.ab,kw,ti OR Suriname.ab,kw,ti OR Syria.ab,kw,ti OR Tajikistan.ab,kw,ti OR Tanzania.ab,kw,ti OR Thailand.ab,kw,ti OR Timor-Leste.ab,kw,ti OR Togo.ab,kw,ti OR Tokelau.ab,kw,ti OR Tonga.ab,kw,ti OR Tunisia.ab,kw,ti OR Turkey.ab,kw,ti OR Turkmenistan.ab,kw,ti OR Tuvalu.ab,kw,ti OR Uganda.ab,kw,ti OR Ukraine.ab,kw,ti OR Uzbekistan.ab,kw,ti OR Vanuatu.ab,kw,ti OR Venezuela.ab,kw,ti OR Vietnam.ab,kw,ti OR (Wallis and Futuna).ab,kw,ti OR (West Bank and Gaza Strip).ab,kw,ti OR Palestine.ab,kw,ti OR Yemen.ab,kw,ti OR Zambia.ab,kw,ti OR Zimbabwe.ab,kw,ti OR Africa.ab,kw,ti OR Asia.ab,kw,ti OR Europe.ab,kw,ti OR America.ab,kw,ti OR Carribean.ab,kw,ti OR Oceania.ab,kw,ti OR low income.ab,kw,ti OR middle income.ab,kw,ti | 1760342 | 21st September 2023 | From Inception till September 21, 2023 |
| 4 | #1 AND #2 AND #3 | 1059 | 21st September 2023 | From Inception till September 21, 2023 |

**Embase**

| SEARCH | SEARCH STRATEGY | RESULTS | DATE OF SEARCH | DATE RANGE |
| --- | --- | --- | --- | --- |
| 1 | Folic.ab,kw,ti. OR folate.ab,kw,ti. OR B9.ab,kw,ti. OR nutrient.ab,kw,ti. OR nutrition.ab,kw,ti. OR fortification.ab,kw,ti. OR policy.ab,kw,ti. OR policies.ab,kw,ti. OR supplement*.ab,kw,ti. OR food.ab,kw,ti. | 1883937 | 21st September 2023 | From Inception till September 21, 2023 |
| 2 | "birth defect*".ab,kw,ti. OR "congenital disorder*".ab,kw,ti. OR "congenital malformation*".ab,kw,ti. OR "congenital abnormalit*".ab,kw,ti. OR "birth anomal*".ab,kw,ti. OR neural tube.ab,kw,ti. OR NTD.ab,kw,ti. OR spina bifida.ab,kw,ti. OR meningocele.ab,kw,ti. OR meningocoele.ab,kw,ti. OR myelomeningocele.ab,kw,ti. OR myelomeningocoele.ab,kw,ti. OR lipomyelomeningocele.ab,kw,ti. OR lipomyelomeningocoele.ab,kw,ti. OR anencephaly.ab,kw,ti. | 90670 | 21st September 2023 | From Inception till September 21, 2023 |
| 3 | LMIC.ab,kw,ti OR (low and middle income countr*).ab,kw,ti OR Afghanistan.ab,kw,ti OR Albania.ab,kw,ti OR Algeria.ab,kw,ti OR Angola.ab,kw,ti OR Argentina.ab,kw,ti OR Armenia.ab,kw,ti OR Azerbaijan.ab,kw,ti OR Bangladesh.ab,kw,ti OR Belarus.ab,kw,ti OR Belize.ab,kw,ti OR Benin.ab,kw,ti OR Bhutan.ab,kw,ti OR Bolivia.ab,kw,ti OR (Bosnia and Herzegovina).ab,kw,ti OR Botswana.ab,kw,ti OR Brazil.ab,kw,ti OR Burkina Faso.ab,kw,ti OR Burundi.ab,kw,ti OR Cabo Verde.ab,kw,ti OR Cape Verde.ab,kw,ti OR Cambodia.ab,kw,ti OR Cameroon.ab,kw,ti OR Cameroun.ab,kw,ti OR Central African Republic.ab,kw,ti OR Chad.ab,kw,ti OR China.ab,kw,ti OR Colombia.ab,kw,ti OR Congo.ab,kw,ti OR Costa Rica.ab,kw,ti OR Cote d'Ivoire.ab,kw,ti OR Ivory Coast.ab,kw,ti OR Cuba.ab,kw,ti OR Djibouti.ab,kw,ti OR Dominica.ab,kw,ti OR Dominican Republic.ab,kw,ti OR Ecuador.ab,kw,ti OR Egypt.ab,kw,ti OR El Salvador.ab,kw,ti OR Equatorial Guinea.ab,kw,ti OR Eritrea.ab,kw,ti OR Eswatini.ab,kw,ti OR Swaziland.ab,kw,ti OR Ethiopia.ab,kw,ti OR Fiji.ab,kw,ti OR Gabon.ab,kw,ti OR Gambia.ab,kw,ti OR Georgia.ab,kw,ti OR Ghana.ab,kw,ti OR Grenada.ab,kw,ti OR Guatemala.ab,kw,ti OR Guinea.ab,kw,ti OR Guinea-Bissau.ab,kw,ti OR Guyana.ab,kw,ti OR Haiti.ab,kw,ti OR Honduras.ab,kw,ti OR India.ab,kw,ti OR Indonesia.ab,kw,ti OR Iran.ab,kw,ti OR Iraq.ab,kw,ti OR Jamaica.ab,kw,ti OR Jordan.ab,kw,ti OR Kazakhstan.ab,kw,ti OR Kenya.ab,kw,ti OR Kiribati.ab,kw,ti OR Korea.ab,kw,ti OR Kosovo.ab,kw,ti OR Kyrgyzstan.ab,kw,ti OR "Lao*".ab,kw,ti OR Lebanon.ab,kw,ti OR Lesotho.ab,kw,ti OR Liberia.ab,kw,ti OR Libya.ab,kw,ti OR North Macedonia.ab,kw,ti OR Madagascar.ab,kw,ti OR Malawi.ab,kw,ti OR Malaysia.ab,kw,ti OR Maldives.ab,kw,ti OR Mali.ab,kw,ti OR Marshall Islands.ab,kw,ti OR Mauritania.ab,kw,ti OR Mauritius.ab,kw,ti OR Mexico.ab,kw,ti OR Micronesia.ab,kw,ti OR Micronesia.ab,kw,ti OR Moldova.ab,kw,ti OR Mongolia.ab,kw,ti OR Montenegro.ab,kw,ti OR Montserrat.ab,kw,ti OR Morocco.ab,kw,ti OR Mozambique.ab,kw,ti OR Myanmar.ab,kw,ti OR Burma.ab,kw,ti OR Namibia.ab,kw,ti OR Nauru.ab,kw,ti OR Nepal.ab,kw,ti OR Nicaragua.ab,kw,ti OR Niger.ab,kw,ti OR Nigeria.ab,kw,ti OR Niue.ab,kw,ti OR Pakistan.ab,kw,ti OR Panama.ab,kw,ti OR Papua New Guinea.ab,kw,ti OR Paraguay.ab,kw,ti OR Peru.ab,kw,ti OR Philippines.ab,kw,ti OR Rwanda.ab,kw,ti OR Saint Helena.ab,kw,ti OR Samoa.ab,kw,ti OR (Sao Tome and Principe).ab,kw,ti OR Senegal.ab,kw,ti OR Serbia.ab,kw,ti OR Sierra Leone.ab,kw,ti OR Solomon Islands.ab,kw,ti OR Somalia.ab,kw,ti OR South Africa.ab,kw,ti OR South Sudan.ab,kw,ti OR Sri Lanka.ab,kw,ti OR Saint Lucia.ab,kw,ti OR (Saint Vincent and the Grenadines).ab,kw,ti OR Sudan.ab,kw,ti OR Suriname.ab,kw,ti OR Syria.ab,kw,ti OR Tajikistan.ab,kw,ti OR Tanzania.ab,kw,ti OR Thailand.ab,kw,ti OR Timor-Leste.ab,kw,ti OR Togo.ab,kw,ti OR Tokelau.ab,kw,ti OR Tonga.ab,kw,ti OR Tunisia.ab,kw,ti OR Turkey.ab,kw,ti OR Turkmenistan.ab,kw,ti OR Tuvalu.ab,kw,ti OR Uganda.ab,kw,ti OR Ukraine.ab,kw,ti OR Uzbekistan.ab,kw,ti OR Vanuatu.ab,kw,ti OR Venezuela.ab,kw,ti OR Vietnam.ab,kw,ti OR (Wallis and Futuna).ab,kw,ti OR (West Bank and Gaza Strip).ab,kw,ti OR Palestine.ab,kw,ti OR Yemen.ab,kw,ti OR Zambia.ab,kw,ti OR Zimbabwe.ab,kw,ti OR Africa.ab,kw,ti OR Asia.ab,kw,ti OR Europe.ab,kw,ti OR America.ab,kw,ti OR Carribean.ab,kw,ti OR Oceania.ab,kw,ti OR low income.ab,kw,ti OR middle income.ab,kw,ti | 2193690 | 21st September 2023 | From Inception till September 21, 2023 |
| 4 | #1 AND #2 AND #3 | 1410 | 21st September 2023 | From Inception till September 21, 2023 |
